# Long-term, ambulatory 12-lead ECG from a single non-standard lead using perceptual reconstruction

**DOI:** 10.64898/2025.12.17.25342224

**Authors:** Sabyasachi Bandyopadhyay, I-Min Chiu, Rayan Ansari, Sheng Liu, J. Weston Hughes, Alexander C. Perino, Neal K. Bhatia, David Ouyang, Euan A. Ashley, Marco V. Perez, James Zou, Sanjiv M. Narayan, Albert J. Rogers

## Abstract

**Background:** Despite its broadening indications, the implantable cardiac monitor (ICM) records a narrow, nonstandard electrocardiogram (ECG) signal which precludes morphological and functional assessments or the application of 12-lead ECG models. We hypothesize that deep learning can be used to reconstruct 12-lead ECG from a single ICM lead for continuously assessing clinical endpoints outside of rhythm detection alone.

**Objective:** To reconstruct 12-lead ECG from a single ICM lead to detect conduction, repolarization, rhythm, and cardiac functional changes in a large, diverse patient population.

**Methods:** We annotated 75,450 echocardiogram-ECG pairs with five disease labels a) right bundle branch block, b) left bundle branch block, c) atrial fibrillation, d) QT-prolongation and e) low left ventricular ejection fraction (LVEF) using regex-based parsing of clinician interpretations. We used perceptual loss to train a deep U-Net (ECG12-PerceptNet) to reconstruct 12-lead ECG from a simulated ICM signal. We compared the classification performance of the reconstructed 12-lead ECG against the original 12-lead and single lead ECG in an internal and external test set. Furthermore, we trained a regression model to predict the absolute LVEF using original and reconstructed 12-lead ECGs.

**Results:** The reconstructed ECG approached the original 12-lead ECG in classification performance across all endpoints while significantly outperforming the single lead ECG. We show two case studies where sequential LVEF measurements were tracked using LVEF predicted with the original and reconstructed 12-lead ECG.

**Conclusion:** In this paper, we report the ECG12-PerceptNet which reconstructs 12-lead ECG from a simulated ICM signal. This can enable continuous in-home or ambulatory monitoring of cardiac functional changes, potentially reducing hospitalizations and out-of-hospital cardiac arrest.

## INTRODUCTION

The electrocardiogram (ECG) is a noninvasive study that is used in the diagnosis of cardiac diseases ranging from myocardial ischemia^1,2^, rhythm abnormalities^3,4^, cardiac structural changes^5^ and arrhythmogenic drugs^6^. Approximately 300 million 12-lead ECGs are performed annually worldwide^7^. Although the 12-lead ECG has been studied for over a century^8^, the recent advance in deep neural networks have greatly expanded the diagnostic capabilities of 12-lead ECGs including ejection fraction^9,10^, wall motion abnormalities^11^, potassium level^12,13^, glucose level^14^, and mortality^15^. Additionally, in recent years, ECG monitoring in the form of wearable patches or implantable cardiac monitors (ICM) with narrow electrode spacing has provided long term continuous ambulatory ECG signal collection capability^16,17^. These monitors have shown a drastic increase in the detection of rhythm abnormalities in patients with cryptogenic stroke^18^ or syncope^19^ over sparsely collected clinical 12-lead ECG measurement or patient-activated devices^20^. However, the non-standard waveforms recorded by these devices do not provide adequate information for monitoring specific disease states such as QT prolongation, interventricular conduction changes, regional ischemia, or disease localization, and do not provide inputs compatible with state-of-the-art deep learning disease classification models.

We set out to provide continuously available 12-lead ECG data to address this unmet need using deep learning. Prior work has applied deep learning techniques to reconstruct 12-lead ECGs from a single standard lead such as lead I^21–24^, lead II^25^, or multiple leads^26–29^. However, these studies have not assessed the output of these models for diagnosing specific clinical endpoints such as QRS morphology or QT prolongation. Additionally, no method has yet been described to reconstruct the 12-lead ECG from a non-standard vector such as that recorded by the ICM. In deep learning, perceptual loss refers to the difference between generated and target images based on high-level features extracted from pretrained convolutional neural networks (CNN), which emphasize perceptual similarities over pixel-level differences^30^. This loss has been widely used in computer vision tasks such as image in-painting, style transfer and super-resolution to make the generated image more visually coherent and realistic to humans. However, this work has not been used to improve the reconstruction quality of time-series signals such as ECGs. We hypothesize that using perceptual loss-based training of a deep neural network we can a) reconstruct 12-lead ECG from a single non-standard ECG lead, b) retain relevant clinical features in the reconstructed 12-lead ECG such that it can robustly identify clinically relevant morphological and rhythm abnormalities and changes in clinical function.

## METHODS

### Study Cohorts

We included data from patients who received a 12-lead ECG at Stanford University between 05/01/2014 and 08/17/2018 and underwent echocardiography within ± 14 days of the ECG measurement. The study was conducted at Stanford University in accordance with the Declaration of Helsinki and was approved by the Stanford University Institutional Review Board (IRB-41045). ECGs were recorded as part of routine clinical care using the Philips iECG application. ECG recordings were ten seconds long at a sampling rate of 500Hz and stored in raw format. Paired echocardiography reports were created by readers with level III American Society of Echocardiography training. **Figure 1** shows the consort diagram for this study. A total of 75,450 echo-ECG pairs were found. We excluded 31,225 pairs when the recorded ECG was not within 14 days of the index echocardiogram and 1,712 where ventricular pacing was involved. Five echo-ECG pairs were excluded due to corrupt ECG files. Finally, we included only the most recent valid echo-ECG pair for each patient, excluding 7,303 multiple examinations. This resulted in 35,210 unique patient cases. An external test dataset was developed using the same criteria at Cedars Sinai with data collected in 2021. 10,610 cases (with paired ECG and echo) met the criteria. **Table 1** includes demographics of the two study populations. Data within the Stanford dataset was stratified into the development cohort consisting of training and validation sets and the hold-out testing cohort as demarcated in **Figure 1**. Stanford University and Cedars Sinai Institutional Review Board approvals were obtained for this study. Waivers of consent form were approved by both IRBs due to the retrospective nature of the study.

**Figure 1.**
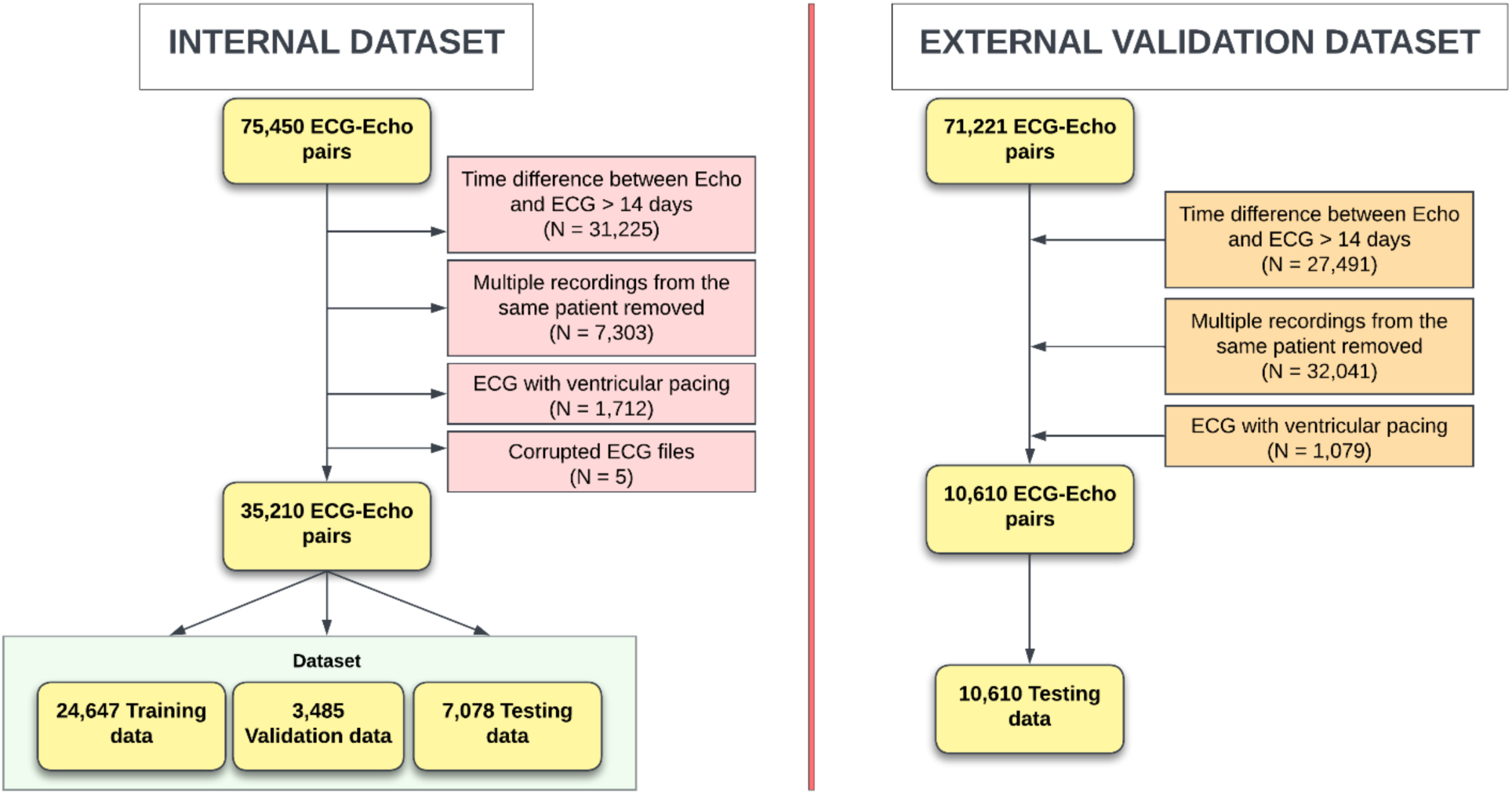
Consensus diagram for creating the Stanford dataset used in this study. Exclusion criteria include time difference between ECG and echo measurement > 14 days, multiple records from the same patient and ECGs with ventricular pacing.

**Table 1.** Demographics of patients in different cohorts.

|  | Training | Validation | Testing | p-value<br>train/valid/test | External<br>validation | p-value<br>test/external |
| --- | --- | --- | --- | --- | --- | --- |
| No. of patients | 15,005 | 3,215 | 6,003 |  | 10,610 |  |
| No. of ECGs | 24,647 | 3,485 | 7,078 |  | 10,610 |  |
| Age (years) | 63.3 ± 17.0 | 63.1 ± 16.9 | 63.8 ± 16.7 | 0.083 | 68.4 ± 16.8 | **0.000** |
| Male | 52.7% | 52.7% | 52.4% | 0.940 | 53.5% |  |
| Race | - | - | - | 0.573 |  | **0.000** |
| Asian | 16.8% | 17.2% | 15.9% | - | 7.4% | - |
| Black | 5.6% | 5.5% | 5.7% | - | 17.6% | - |
| Native American | 0.4% | 0.4% | 0.4% | - | 0.4% | - |
| Other | 16.3% | 17.0% | 16.9% | - | 7.5% | - |
| Pacific Islander | 1.2% | 1.5% | 1.1% | - | 0.4% | - |
| Unknown | 4.7% | 4.5% | 5.0% | - | 2.4% | - |
| White | 55.1% | 53.9% | 55.0% | - | 64.3% | - |
| Ethnicity | - | - | - | 0.769 |  | **0.000** |
| Hispanic/Latino | 11.1% | 11.4% | 11.2% | - | 12.9% | - |
| Non-Hispanic | 82.7% | 82.6% | 82.2% | - | 84.3% | - |
| Unknown | 6.3% | 6.0% | 6.6% | - | 2.8% | - |
| HTN | 63.0% | 62.2% | 63.0% | 0.670 | 61.8% | **0.000** |
| HF | 27.3% | 27.5% | 27.4% | 0.939 | 32.5% | **0.000** |
| AF | 9.3% | 9.4% | 9.5% | 0.300 | 9.8% | **0.000** |
| RBBB | 9.3% | 9.1% | 9.1% | 1.000 | 11.3% | **0.000** |
| LBBB | 4.5% | 4.5% | 4.3% | 1.000 | 3.5% | **0.000** |
| Prolonged QT | 5.7% | 6.0% | 5.7% | 1.000 | 6.0% | **0.000** |
| Echocardiography |  |  |  |  |  |  |
| WMA | 10.9% | 9.8% | 10.6% | 0.161 | 1.9% | **0.000** |
| LVEF | 58.9 ± 11.2 | 58.8 ± 11.5 | 59.0 ± 11.0 | 0.637 | 58.5 ± 13.9 | **0.009** |
| Low LVEF | 5.5% | 6.2% | 5.0% | **0.000** | 8.1% | **0.000** |

Abbreviations. AF; Atrial Fibrillation, ECG; Electrocardiogram, HF; Heart failure, HTN; Hypertension, WMA; Wall Motion Abnormality, LBBB; Left Bundle Branch Block, RBBB; Right Bundle Branch Block, PVC; Premature Ventricular Complexes, LVEF; Left Ventricular Ejection Fraction.

ICM device-recordings (10 secs long, sampled at 256 Hz) with interrogation between 2016 and 2025 were extracted using a custom function from PDF reports in 56 patients. Among these, 33 patients had at least one 12-lead ECG within 1 month of the device recording. If multiple 12-lead ECGs were available within 1 month, the closest one to the ICM recording was chosen as the “comparator”. This cohort (N=33) was used to evaluate the ICM to V3 – V2 similarity and inter V3 – V2 similarity to demonstrate the equivalence between using a real ICM versus V3 – V2 signal from the clinical 12-lead ECG, for the purpose of reconstructing the complete 12-lead ECG.

### Clinical Endpoints

Clinical endpoints included both ECG and echocardiogram derived cardiac abnormalities. We parsed interpretations of 12-lead ECGs provided by board-certified cardiologists and electrophysiologists during routine clinical care to create labels for cardiac abnormalities. In order to assess abnormalities of rhythm, depolarization (morphology) and repolarization ECG labels were parsed from verified interpretations, using positive and negative search terms in a nested loop (**Supplementary Table 1**). The following conditions were identified from the textual interpretations: a) Right Bundle Branch Block (RBBB), b) Left Bundle Branch Block (LBBB), c) QT-segment prolongation, d) Atrial Fibrillation (AF). Interpretations in text format were a) mined for positive terms indicating the presence of each of these abnormalities and b) mined for negative words and phrases which indicated the absence/resolution of abnormalities.

The Left Ventricular Ejection Fraction (LVEF) was assessed using paired echocardiograms within 14 days of the ECG. This endpoint was used to assess the ability to predict changes in cardiac function. LVEF was parsed from semi-structured echocardiographic reports. The Low LVEF label corresponded to LVEF <= 35.

### 12-lead ECG Reconstruction

We trained a U-Net model called ECG12-PerceptNet to reconstruct the 12-lead ECG signal from a single-lead narrowly spaced ECG waveform corresponding to the recording vector of an ICM. This representative signal was derived from V3 – V2. We simultaneously trained another U-Net generator to reconstruct the V3-V2 signal from itself. These two U-Net generators shared the same decoder, and their latent vectors were coupled during training using a regularization loss. Furthermore, to improve the reconstruction of clinically relevant features, we regularized the U-Net generator used for reconstructing the 12-lead ECG with a perceptual loss generated from a classifier network which was pretrained on our training dataset with the five disease labels. The final loss function includes a) image reconstruction loss, b) perceptual loss, c) latent vector loss and d) total variance reduction loss. The latent vector loss ensures that the 12-lead reconstruction does not use features other than those available from the single-lead ECG. The total variance loss constrains the total variance of the output signal which prevents overfitting to the training data distribution. The final loss function is mathematically represented by the following equation:

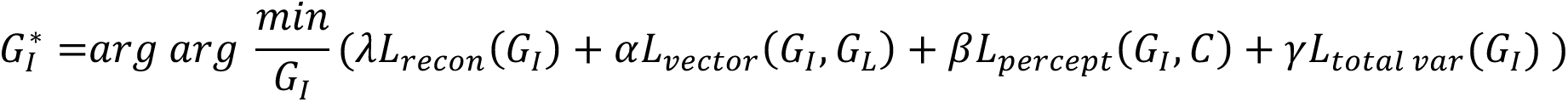

Here *G_I_* represents the generator used to reconstruct the 12-lead ECG signal, 𝐺_*L*_ represents the parallel generator reconstructing the single lead (V3-V2) ECG signal, 𝐶 represents the pretrained 12-lead ECG classifier which is used to create the perceptual loss. Training was conducted for 10 epochs, with a batch size of 64 and learning rate of 0.001. Learning rate scheduling did not significantly improve performance on the validation set. Optimal values for the coefficients of the different terms inside the loss function were selected by performing a grid search. The best values of 𝜆, 𝛼, 𝛽, 𝛾 were found to be 25, 0.5, 250 and 5e-06 respectively. The third hidden layer of the classification network was used to create perceptual loss. This was chosen through grid search. The performance of the third hidden layer was identical to the performance from the entire classifier hence the third layer was chosen for faster computation at runtime. Python v3.7 and Tensorflow v2.15.0 were used to develop the model. **Figure 2** shows the architecture of the ECG12-PerceptNet + 12-lead classifier. We used the following metrics for quantifying the lead-wise similarity between original and reconstructed 12-lead signals: Mean Squared Error (MSE), Mean Absolute Error (MAE), Maximum Mean Discrepancy (MMD) and Structural Similarity Index (SSIM). We compared the reconstruction performance against the EK-GAN architecture published by Joo et.al., 2023^23^.

**Figure 2.**
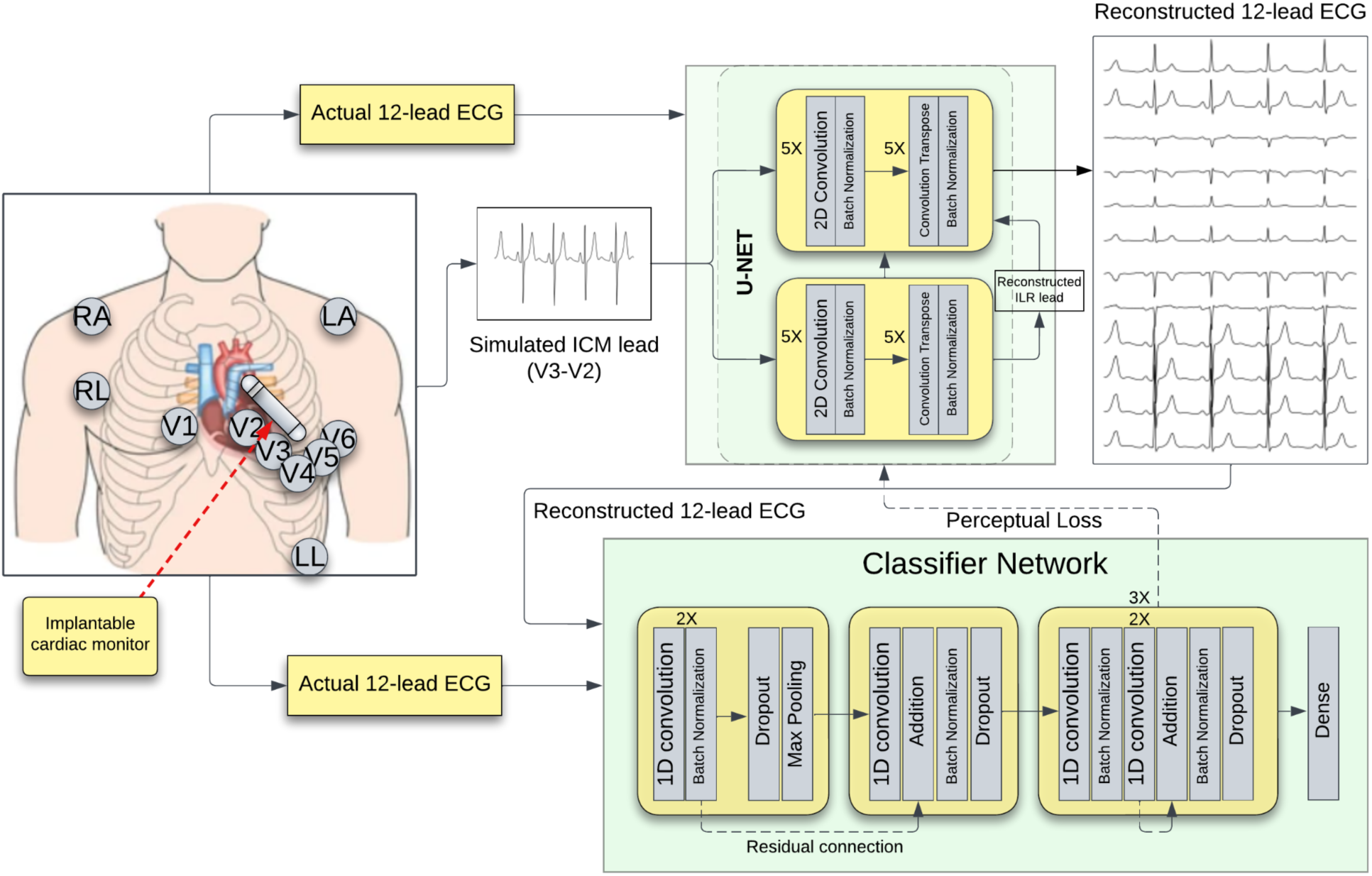
Combined architecture of the U-Net perceptual loss network for reconstructing 12-lead ECG from single lead and the pretrained 12-lead ECG classification network. Differences between the actual 12-lead ECG and the reconstructed 12-lead ECG is used to train the reconstruction network. Additionally, differences between the latent embeddings generated by the actual 12-lead and the reconstructed 12-lead signals through the pretrained classifier are used as a perceptual loss for refining the reconstruction network.

### Model Performance

We used the ECG12-PerceptNet generator + pretrained classifier combination to evaluate classification performance of the reconstructed 12-lead ECG across the five clinical endpoints. We compared this against the classification performances of the original 12-lead ECG and the original single lead ECG signals using the same 12-lead classifier. Furthermore, we used the same classifier to compare the performances of the 12-lead ECG reconstructed using the ECG12-PerceptNet generator against that reconstructed using the EK-GAN generator. Finally, we compared the classification performance of the reconstructed 12-lead ECG signals using the 12-lead classifier against the single lead ECG using a dedicated single lead classifier trained on the training dataset. This single-lead classifier had a ResNet architecture similar to the 12-lead classifier. We used a combination of the following metrics to assess performance: area under receiver operating curve (AUROC), accuracy, average precision, F1-score, sensitivity, specificity, and negative predictive value (NPV). The classification scores were bootstrapped 1000 times with replacement to generate 95% confidence intervals (CI) for statistically comparing different models. An identical methodology was used to assess the classification performances using the external validation dataset.

### LVEF Regression and Clinical Transition Detection

We adapted the 12-lead ECG classifier to a regression network of identical architecture, replacing the classification head with a single linear output and fine-tuning it using the absolute LVEF values from the training dataset. The fine-tuning was performed using the Adam optimizer (learning rate=1e-3) and Huber Loss (delta=0.08) with learning rate reduction on validation loss plateaus and early stopping using the validation loss. The model was simultaneously trained on two data sources-a) actual 12-lead ECG, and b) reconstructed 12-lead ECG from ECG12-PerceptNet. We introduced source-awareness without changing the network backbone by incorporating a FiLM (feature wise linear modulation) layer between the convolutional stem and the residual stack of the network. A binary domain token (0: original ECG, 1: reconstructed ECG) was applied to the FiLM layer which introduced per-channel scaling/shifts in the feature map before the residual stack. We also conducted post-hoc, domain-specific calibration on the validation set, wherein we calculated percentiles from the ground-truth LVEF and predictions, and used a piece-wise linear mapping function to map each prediction to its target quantile.

We also used the predictions to classify LVEF drop/recovery events between consecutive echo pairs per domain. We converted the predictions to a continuous crossing strength score defined by the following equations. For each consecutive visit pair (*y_prev_, y_next_*) and threshold 𝑇 = *35* we computed

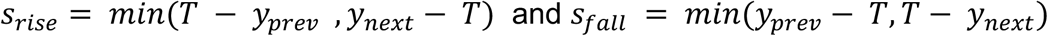

A true transition across the threshold (LVEF=35) would make the score positive, while a non-event would make the score negative, thereby simplifying the classification problem to one of sign detection. We reported the AUROC, Sensitivity, Specificity, Recall and NPV with 95% confidence intervals obtained using patient-wise resampling 2000 times.

### Statistical Methods

The following statistical methods were used in this study: for comparing clinical demographics, unpaired two-tailed Student’s t-test was used for continuous variables and the Chi-squared test was used for categorical variables. For continuous variables, normality was assessed using the Shapiro-Wilk test, and non-normal variables were compared using the Mann-Whitney U Test. Continuous variables are reported as mean ± S.D. and categorical variables are reported as percentages. AUROC, Average Precision, Accuracy, F1-Score, Sensitivity, Specificity and NPV are reported for assessing multi-disease classification by the deep learning models such as ECG12-PerceptNet. Prediction scores were bootstrapped 1000 times with replacement to generate 95% confidence intervals.

## RESULTS

### Participants

The Stanford dataset consists of a roughly equal percentage of male and female patients with an average age of 63 years and 45% data from minority populations (**Table 1**). Two-thirds of these patients suffered from hypertension; a third of the patients had a previous history of heart failure and one-tenth of patients suffered from cardiac wall motion abnormalities. The incidence rates of the cardiac abnormalities classified in this study were as follows: 9% for RBBB, 4% for LBBB, 6% for QT-prolongation, 9% for AF and 5% for low LVEF. There were no statistically significant differences between the prevalence of these abnormalities across the training, validation and testing cohorts in the Stanford dataset (**Table 1**). The external validation dataset contained significantly older patients compared to the Stanford dataset with the average age being 68 years and had a significantly higher percentage of data from the African American population. Patients in this dataset had a significantly lower prevalence of cardiac wall motion abnormalities but higher prevalence of low LVEF (**Table 1**). **Table 1** lists the demographics for the two study populations.

### 12-lead ECG Reconstruction

The ECG12-PerceptNet generated 12-lead ECG reconstructions with significantly improved MAE, MSE and SSIM across all leads compared to the EK-GAN generator network. The ECG12-PerceptNet and EK-GAN generators showed comparable MMD values across all leads except lead III. **Supplementary Table 2** shows all reconstruction error values from the ECG12-PerceptNet and EK-GAN.

### Equivalence between ICM and V3 – V2

A cohort of 33 patients were used to compare the actual ICM recordings versus V3 – V2 extracted from 12-lead ECGs with respect to the initial single lead waveforms and the 12-lead reconstructions generated from those signals by ECG12-PerceptNet (**Figure 3**). The V3 – V2 signal nearest to the ICM recording was used as the comparator signal across all panels. **Figure 3A** highlights multiple serial V3 – V2 signals (**blue**) which show substantial variability possibly due to variation in the location of the precordial leads during routine 12-lead ECG acquisition and the ICM signal collected from the same patient (**green**). **Figure 3B** shows a comparison of the original 12-lead ECG (**red**) adjacent in time to the ICM recording with the reconstructed 12-lead ECG from both the ICM recording (**green**) and the V3-V2 signal from serial ECGs (**blue**). **Figure 3C** shows that MSE differences between the single lead tracings (ICM or V3-V2) with corresponding temporally adjacent single lead recording (V3-V2) are greater than the MSE differences between their respective 12-lead reconstructions. This shows that the reconstruction network effectively acts as a rectifier for spurious variations while retaining clinically meaningful signals as demonstrated by the classification performances. However, this difference in MSE comparisons between temporally adjacent single lead waveforms or 12-lead reconstructions was not statistically significant after correction for multiple comparisons.

**Figure 3.**
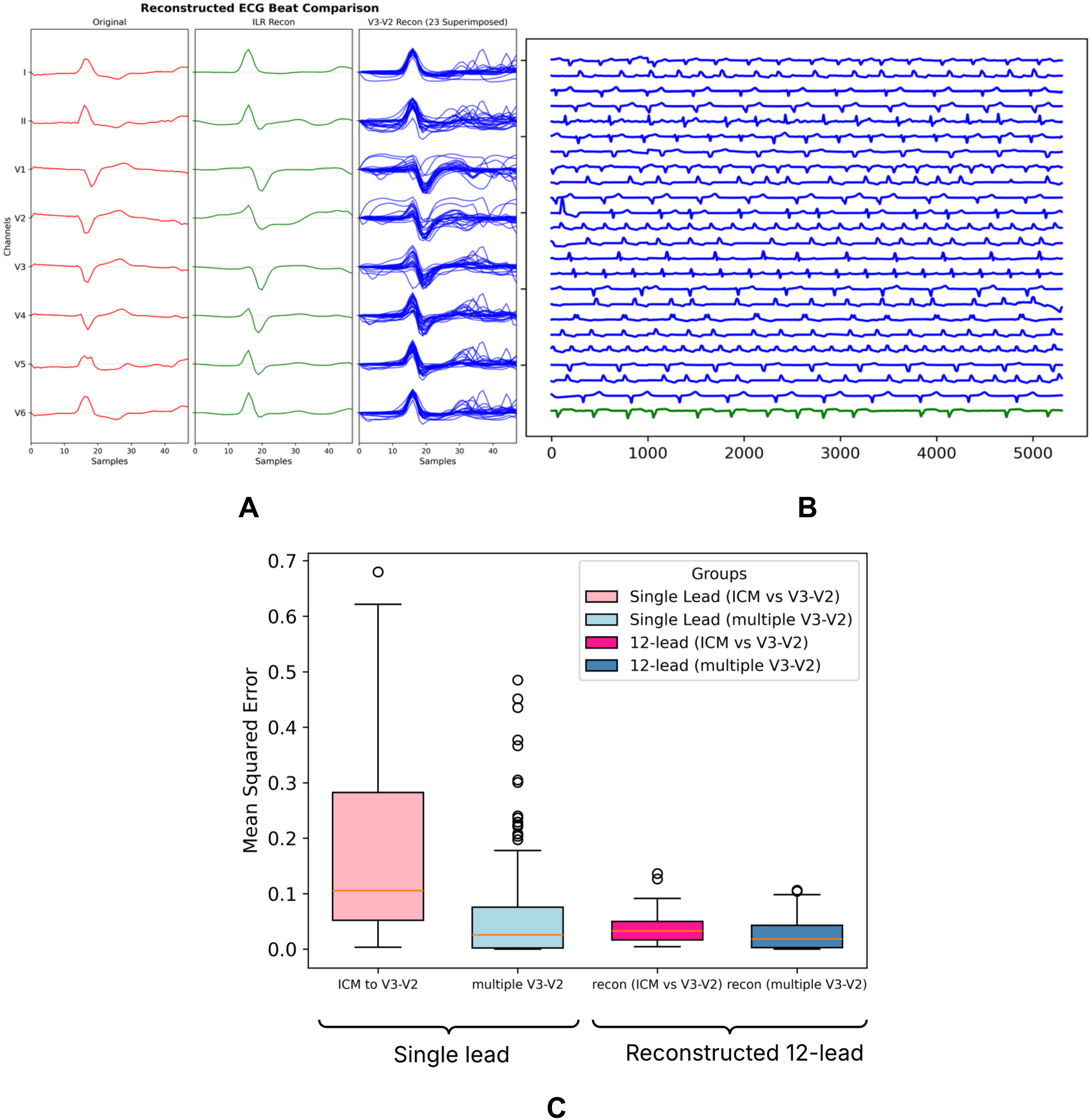
ICM signal and V3-V2 signals are comparable. Reconstructed 12-lead ECG from original ICM and from V3-V2 are similar. A) Comparison of beat morphologies between original 12-lead ECG, reconstructed 12-lead ECG from ICM signal and reconstructed 12-lead ECG from V3-V2 signals collected within 1 month of the ICM recording for a randomly selected patient. Beat morphologies reconstructed from different V3-V2 signals from the same patient (within 1 month) exhibit sufficient heterogeneity broad enough to encompass the morphology derived from the ICM-based 12-lead reconstruction. B) Visual comparison between 10-second recordings of V3-V2 (in blue) and actual ICM recording (in green, bottom) shows that V3-V2 signals from the same patient exhibit sufficient heterogeneity broad enough to encompass the reference ICM template. C) Mean squared errors between ICM and nearest (in time) V3-V2 (comparator), comparator and other V3-V2 from same patient, reconstructed 12-lead ECG from ICM versus that from comparator, and reconstructed 12-lead ECG from comparator versus those from other V3-V2 from same patient. The boxplots are not significantly different (adjusted p > 0.05) when accounting for multiple comparisons. The difference between the reconstructed 12-lead ECG is smaller than the difference between the single channel waveforms (ICM or V3-V2).

### Model Performance

The ECG12-PerceptNet generated 12-lead ECG signals outperformed the original single-lead ECG and approached the performance of the original 12-lead ECG signal across all clinical endpoints in both the internal hold-out test and the external validation cohorts. **Figure 4** shows the AUROC curves for classifying LBBB, AF and LVEF using the original 12-lead ECG, reconstructed 12-lead ECG and original single lead ECG signals. The ECG12-PerceptNet generated 12-lead ECG outperformed the EK-GAN generated 12-lead ECG in classifying all clinical endpoints. **Supplementary Figure 1** shows the AUROC curves comparing the ECG12-PerceptNet and the EK-GAN reconstructions in LBBB, AF and LVEF. Finally, the ECG12-PerceptNet reconstructed 12-lead ECG outperformed the original single lead ECG when a dedicated single lead classifier was used (**Supplementary Fig 2**). **Supplementary Table 3** and **Supplementary Table 4** show all performance metrics across all clinical endpoints in the internal hold-out test dataset and the external validation dataset respectively.

**Figure 4.**
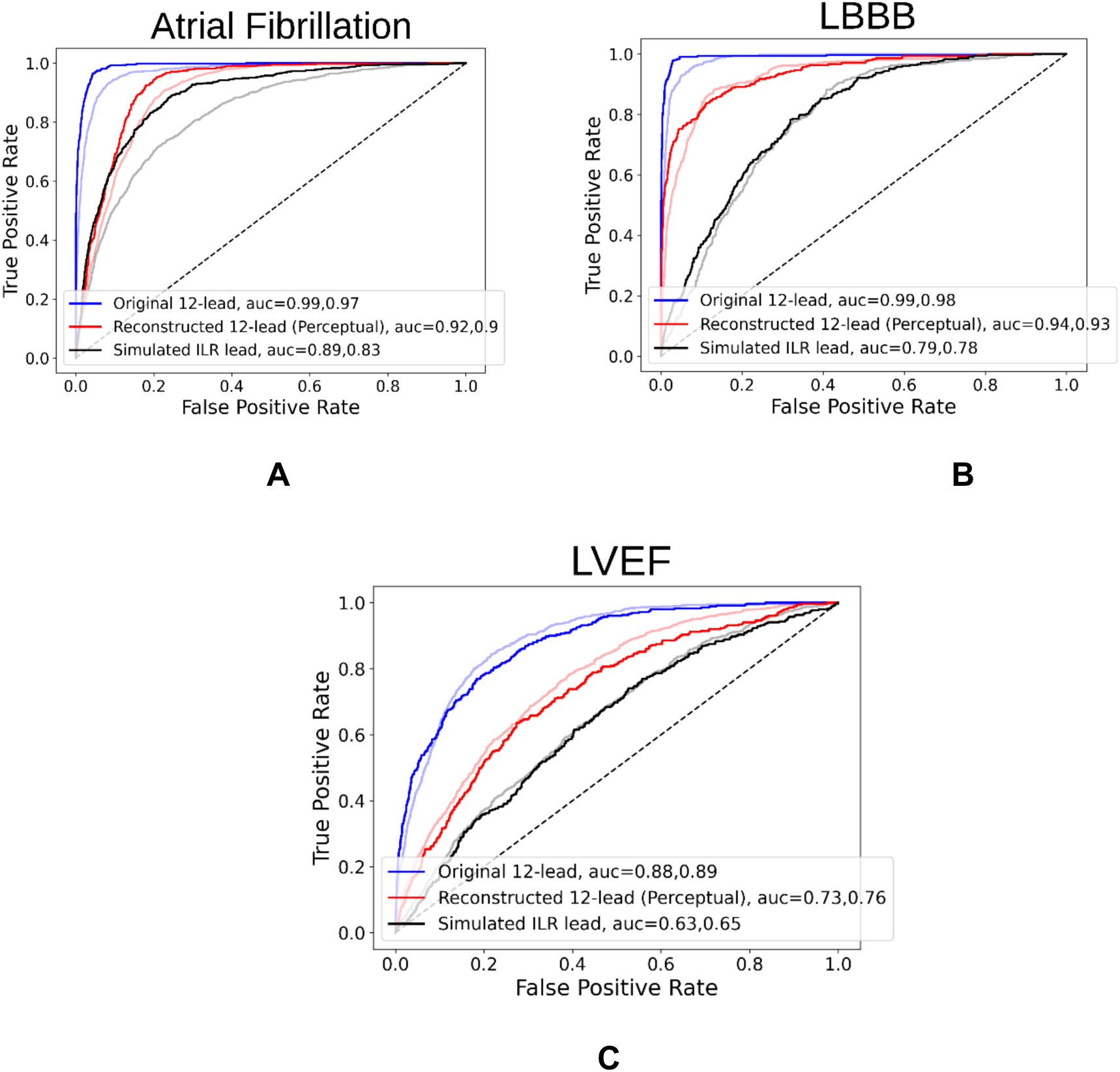
A comparison between the area under receiver operating curve (AUROC) values between the original 12-lead ECG, reconstructed 12-lead ECG and single lead ECG signals for a rhythm abnormality (Atrial fibrillation; panel A), morphological abnormality (left bundle branch block; panel B) and functional abnormality (low ejection fraction; panel C). The performances in the Stanford dataset (solid lines) are overlaid on the performances in the independent external validation dataset (faded lines) for comparison.

### LVEF Regression and Clinical Transitions Classification

We predicted the absolute LVEF values using a regression model which was fine-tuned for the LVEF regression task using our training dataset. We used both the original 12-lead ECG and the ECG12-PerceptNet reconstructed 12-lead ECGs to predict the absolute LVEF over sequential recordings. The FiLM-conditioned LVEF regression model with per-domain quantile calibration achieved a correlation coefficient of 0.63 between predicted and reference LVEF for original 12-lead ECG and 0.46 for reconstructed 12-lead ECG. The reconstructed signals showed a narrower prediction spread, which was corrected at inference using the quantile calibration which eliminated distributional mismatch between the two domains. On the LVEF drop and recovery classification task with reference to LVEF = 35, the model showed discriminative ability for both original and reconstructed ECGs. For original 12-lead ECG, AUC = 0.78 (95% CI 0.75-0.81), sensitivity = 0.82 (CI 0.72-0.89), specificity = 0.63 (CI 0.54-0.72), NPV = 0.99 (CI 0.99-1.00) for LVEF drops, and AUC = 0.76 (CI 0.74-0.78), sensitivity = 0.85 (CI 0.62-0.91), specificity = 0.53 (CI 0.49-0.75), NPV = 0.99 (CI 0.99-1.00) for LVEF recoveries. For reconstructed 12-lead ECG, AUC = 0.71 (CI 0.68-0.74), sensitivity = 0.70 (CI 0.58-0.87), specificity = 0.61 (CI 0.45-0.73), NPV = 0.99 (CI 0.99-1.00) for LVEF drops, and AUC = 0.71 (CI 0.69-0.74), sensitivity = 0.68 (CI 0.59-0.82), specificity = 0.65 (CI 0.51-0.72), NPV = 0.99 (CI 0.99-1.00) for LVEF recoveries. **Figure 5** shows the ROC curves for drop/recovery classification from the original and reconstructed 12-lead ECGs.

**Figure 5.**
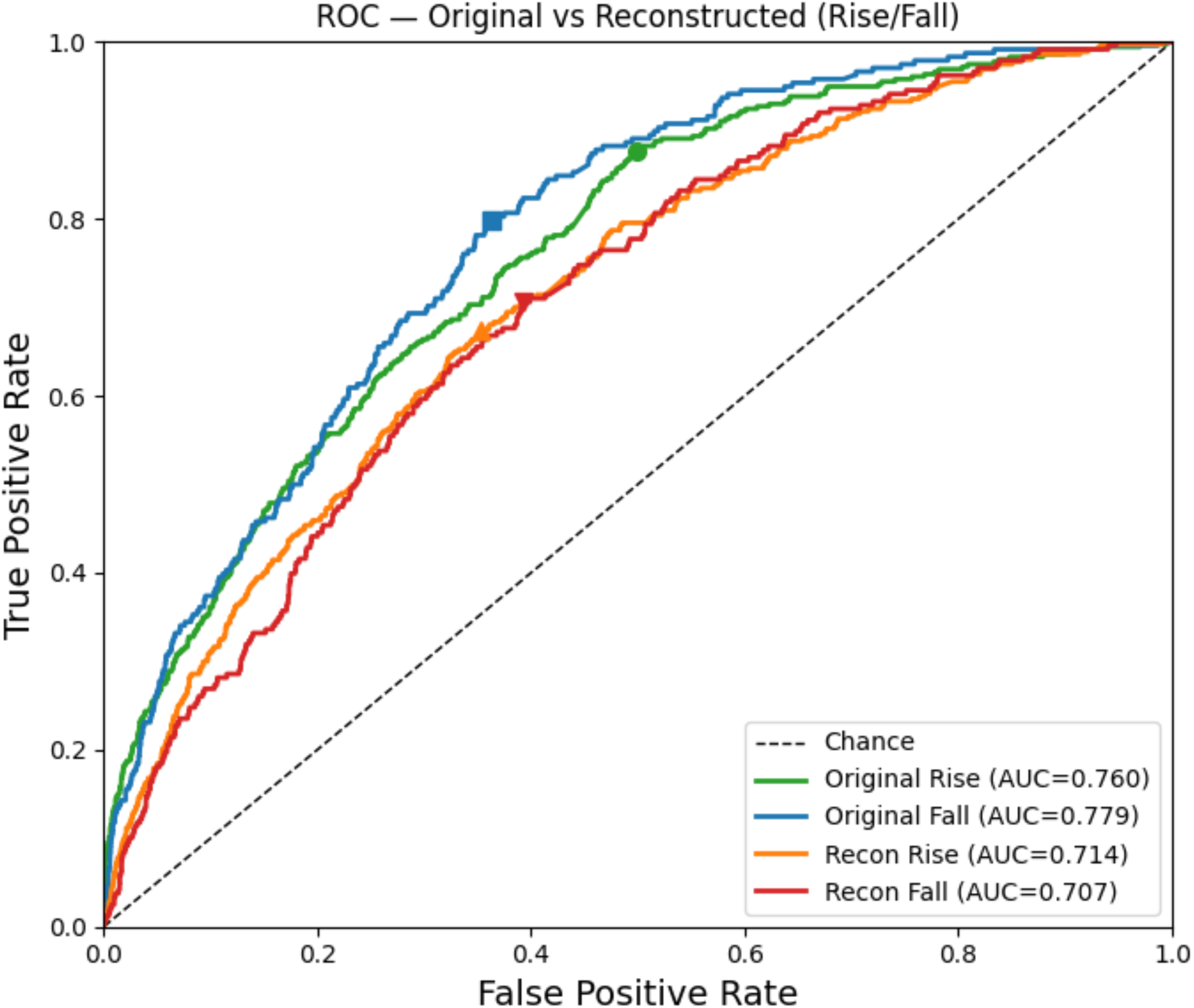
Receiver Operating Characteristics (ROC) showing the performance in classifying LVEF drop and rise events across the predetermined threshold (LVEF = 35) using the original 12-lead ECGs (drop:blue/rise:green) and reconstructed 12-lead ECGs (drop:red/rise:orange) respectively.

## DISCUSSION

In this paper, we present ECG12-PerceptNet, a U-Net generator which reconstructs a standard 12-lead ECG from a single non-standard precordial ECG recording vector. This model expands continuous monitoring capabilities to include morphological, functional and conduction abnormalities. Our approach incorporates features vital to clinical classifications through perceptual loss, thus providing state-of-the-art performance. We have validated the performance of the reconstructed 12-lead ECG signals in classifying rhythm abnormalities (AF), morphological abnormalities (RBBB/LBBB/QT prolongation) and cardiac function (LVEF). The ECG12-PerceptNet model has provided greatly improved rhythm, morphology and function classification over the single-lead baseline while using both the standard 12-lead disease classifier and a single lead disease classifier. The large and diverse Stanford dataset resulted in robust performance in an external validation dataset from a demographically distinct population. The ECG12-PerceptNet improved the reconstruction quality over previous approaches^23^. Finally, we showed that the 12-lead reconstructions from real ICM signals were similar to those from V3 - V2, despite considerable variability in serial V3 - V2 recordings. MSE calculations showed that the ECG12-PerceptNet reconstructions had significantly lower variance than that between corresponding single lead waveforms. This highlights the ability of the ECG12-PerceptNet to generalize robust reconstructions from our V3 - V2 training dataset and across ICMs despite variability in the implant location.

There are significant clinical implications of our study. Currently, long term monitoring is limited by the use of nonstandard, narrowly spaced electrodes in the ICM. Alternative non-continuous wearable options require patient activation and compliance. This prevents measurement during debilitating arrhythmia episodes where the patient might suffer from syncope or transient arrhythmias.^31^ An accurate continuous long-term 12 lead monitoring solution would provide improved ambulatory screening of patients at risk of cardiac failure, rhythm abnormalities, QT prolongation, regional ischemia, or sudden cardiac death. Furthermore, conversion of single-lead to 12-lead ECGs permits widescale use of previously developed 12-lead classifiers for ambulatory monitoring of a host of other endpoints such as potassium levels^13^, glucose levels^32^, mortality risk or preeclampsia^33^. In this study, we found that the model identified clinically important LVEF depression and compensation events in patients with echocardiogram-electrocardiogram pairs from a single lead with similar performance as the original 12-lead ECG.

Our study has limitations. The model was trained on simulated ICM signals (V3 - V2) due to the inability to obtain a large-scale database of simultaneous ICM and 12-lead ECGs. In the absence of such a registry, we created ICM-like waveforms from V3-V2 of the 12-lead ECG to train our foundational reconstruction model, and showed the near equivalence of reconstructions from true ICM signals and V3-V2 in patients with implants. The training dataset is based on 10 second strips in supine patients rather than continuous data, although the ICM recordings in this study are random ‘presenting rhythms’ of device data downloads in the ambulatory setting. Finally, clinical thresholds for ambulatory alerts are yet to be determined and may vary based on the disease being monitored.

Future work will focus on expanding the reconstruction and classification network to individual beat-level instead of ten seconds fragments. This might improve the reconstruction and classification performances of transient arrhythmias such as premature ventricular contractions of different morphologies or regional ischemia. The current reconstruction paradigm can be further expanded to reconstruct high density lead arrangements such as electrocardiographic imaging^34^ from 12-lead ECG to enable real-time arrhythmia source localization. Finally, miniaturization of the neural network models using techniques such as convex polishing^35^ can enable point-of-care deployment of these models. This will help reduce prediction latency which can be beneficial for applications such as real-time monitoring of the risk of sudden cardiac arrest.

## Summary

This study provides a model and method to reconstruct all 12 standard ECG leads from a non-standard narrowly spaced precordial ECG recorder. We show that V3 – V2 recordings extracted from clinical 12-lead ECGs are computationally equivalent to original ICM recordings. Additionally, after reconstruction of the full lead set, the reconstructed 12-lead ECGs are even more closely matched than the original single lead signals. We improved the classification performance of the reconstructed 12-lead ECG by utilizing classification features as an additional reconstruction loss through perceptual training. Finally, we created an LVEF regression network by transferring the weights of the classification model and fine-tuning using LVEF data to demonstrate its ability to detect LVEF drop/rise events. Future work will expand to panoptic reconstruction to generate dense ECGi waveforms from a single-lead ICM for performing regional arrhythmia source localization.

## OTHER INFORMATION

### Data availability

The raw patient data are not publicly available due to institutional policy and human subjects’ approval to protect patient privacy. For completely deidentified data please contact A.J.R. at. Data can be shared upon reasonable request.

### Code availability

The source code for this project, including ECG12-PerceptNet model architecture and language processing code is available on GitHub at https://github.com/sabyasachi0212/ECG-reconstruction-using-perceptual-loss.git

### Ethics Declarations

This study was approved by the Stanford University Institutional Review Board (IRB 41045) and the Cedars-Sinai Institutional Review Board (IRB Study 1506). The requirement for informed consent was waived by both IRBs because the study involved retrospective analysis of fully de-identified clinical data which presented minimal risk to participants. All research procedures were performed in accordance with relevant guidelines and regulations, including the Declaration of Helsinki.

## Contributions

A.J.R., R.A., and S.B. conceived of the study. S.B., J.W.H., R.A., and A.J.R. were involved in acquiring the Stanford dataset in a format appropriate for this study and preprocessing. I.M.C., and D.O. performed data acquisition, preparation, and analysis at Cedars Sinai Medical Center. S.B., R.A., A.J.R. performed data analysis at Stanford. S.B., S.L., and A.J.R. performed model development and optimization. The work was led by S.B., and overseen by N.B., E.A.A., M.V.P., D.O., J.Z., S.M.N., A.C.P and A.J.R. All authors provided comments and approved the manuscript.

## List of Abbreviations

AF: atrial fibrillation
AUROC: area under the receiver operating curve
CI: confidence interval
ECG: electrocardiogram
GAN: generative adversarial network
ICM: implantable cardiac monitor
LBBB: left bundle branch block
LVEF: left ventricular ejection fraction
MAE: mean absolute error
MMD: maximum mean discrepancy
MSE: mean squared error
NPV: negative predictive value
RBBB: right bundle branch block
SSIM: structural similarity.

