## Supplemental Materials for "Long-term, ambulatory 12-lead ECG from a single non-standard lead using perceptual reconstruction"

SUPPLEMENTARY INFORMATION

This document contains supplementary information for the manuscript Bandyopadhyay et.al., “Long-term, ambulatory 12-lead ECG from a single non-standard lead using perceptual reconstruction”, 2025.

### Supplementary Figures

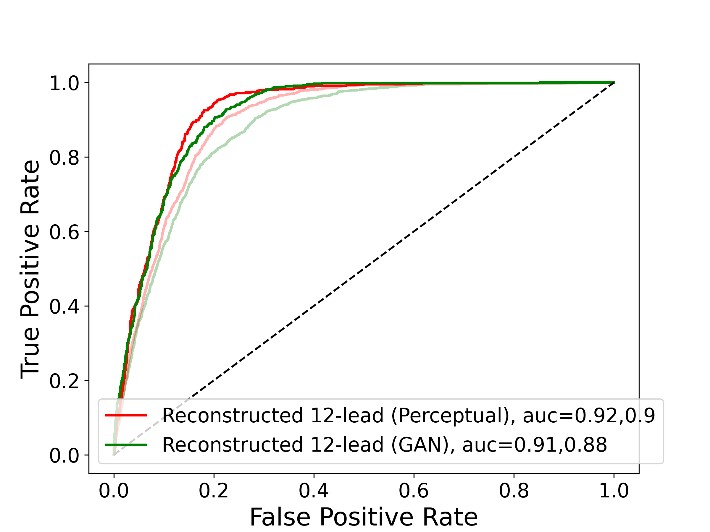

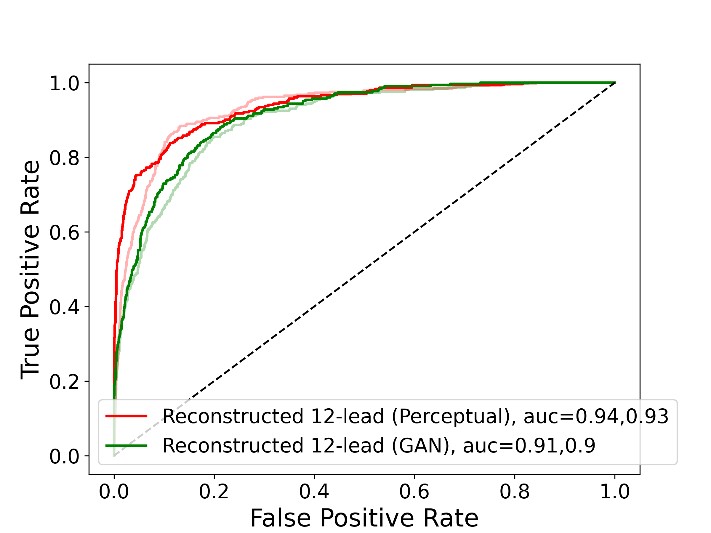

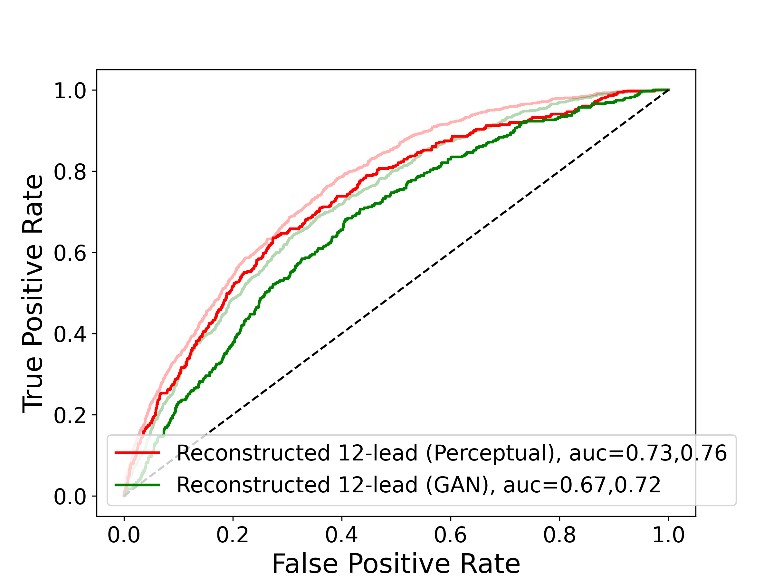

**Supplementary Figure 1.** A comparison between the area under receiver operating curve (AUROC) values between the reconstructed 12-lead ECG constructed using the ECG12-PerceptNet network (red) and GAN network (green) for a rhythm abnormality

(Atrial fibrillation; panel A), morphological abnormality (left bundle branch block; panel

B) and functional abnormality (low ejection fraction; panel C). The performances in the The performances in the Stanford dataset (solid lines) are overlaid on the performances in the independent external validation dataset (faded lines) for comparison.

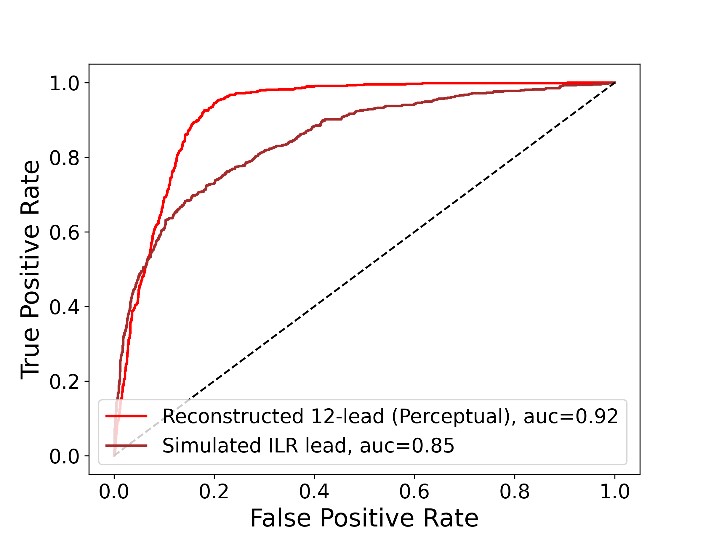

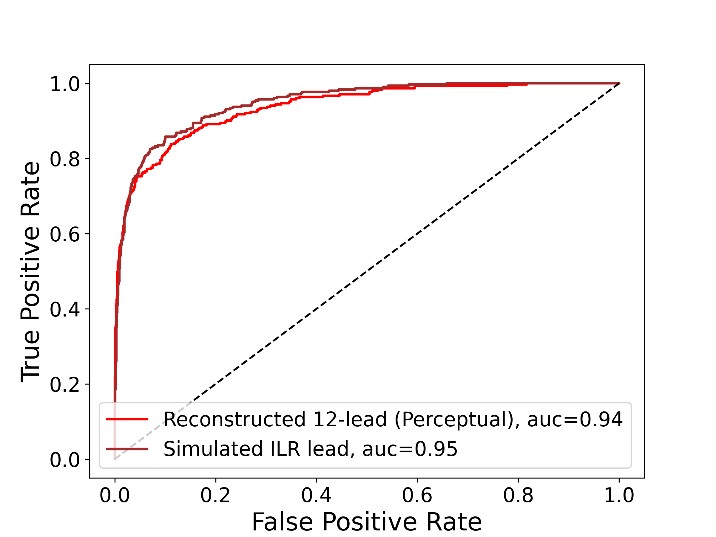

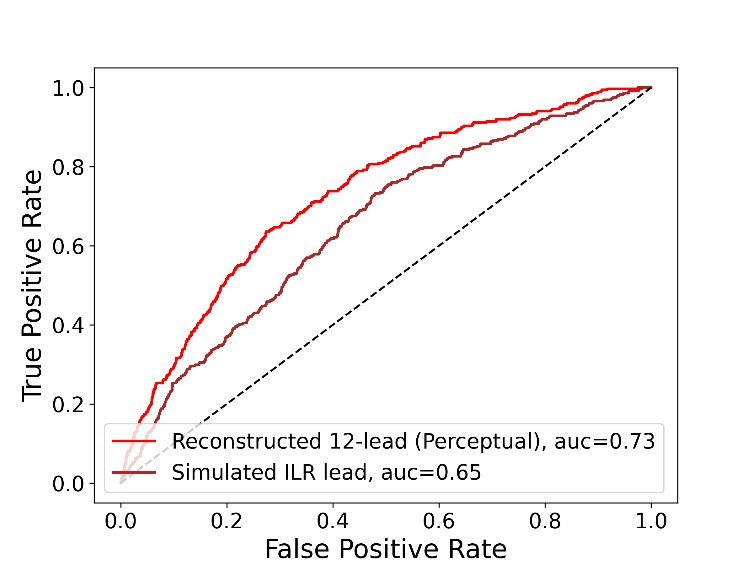

**Supplementary Figure 2.** A comparison between the area under receiver operating curves (AUROC) of the reconstructed 12-lead ECG classified using the pre-trained 12lead classifier (red) versus the single lead ECG (V3-V2) classified using a dedicated single-lead for a rhythm abnormality (Atrial fibrillation; panel A), morphological

abnormality (left bundle branch block; panel B) and functional abnormality (low ejection fraction; panel C).

### Supplementary Tables

**Supplementary Table 1.** Positive and Negative expressions for creating disease labels.

|  | Positive expressions | Negative expressions |
| --- | --- | --- |
| Right bundle branch block | 'RIGHT BUNDLE BRANCH  BLOCK', 'RBBB' | 'HAS DISAPPEARED', 'NO  LONGER PRESENT', 'NO  LONGER PRESENT', 'IS NOT  PRESENT', 'NO RBBB', 'RBBB  NO LONGER SEEN',  'DISAPPERANCE OF RIGHT  BUNDLE BRANCH BLOCK',  '[NOW ABSENT]', 'NO  LONGER EVIDENT', 'NO  LONGER EVIDENT', 'RIGHT  BUNDLE BRANCH BLOCK  WAS PRESENT ON THE  PRIOR EKG', 'IS NOT  EVIDENT TODAY', 'A  CLASSICAL PATTERN WAS  PRESENT ON THE PRIOR  EKG', ', WAS RBBB', 'RBBB  AND LAFB PATTERN WAS  PRESENT ON THE PRIOR  EKG', 'PATTERN HAS  CHANGED FROM RBBB',  'PRIOR LPFB/IRBBB', 'RBBB  CONVERTED' |
| Left bundle branch block | 'LEFT BUNDLE BRANCH  BLOCK', 'LBBB' | 'HAS DISAPPEARED', 'NO  LONGER PRESENT', 'IS NOT  PRESENT', 'IS NOT  PRESENT NOW', 'IS NOW  ABSENT', 'IS NO LONGER  PRESENT', 'HERE IS NO  LBBB', '[NOW ABSENT]', 'NO  LONGER EVIDENT', 'DO NOT  HAVE AN LBBB PATTERN',  'CRITERIA FOR LBBB ARE NO  LONGER MET', 'IS DIFFERENT  FROM PRIOR LBBB',  'CHANGED FROM LBBB',  'LEFT BUNDLE BRANCH  BLOCK WAS PRESENT  ON THE PRIOR EKG', 'IS PRESENT ON PRIOR  EKG', 'IS PRESENT IN ALL  PRIOR EKGS', 'HAD A NEW  LBBB PATTERN', 'PRIOR  LBBB', 'DOES NOT HAVE  AN LBBB PATTERN', 'DO |
|  |  | NOT HAVE AN LBBB  PATTERN', 'LBBB PATTERN  WAS PRESENT ON A  PRIOR EKG' |
| Atrial fibrillation | 'ATRIAL FIBRILLATION',  'ATRIAL  FLUTTER/FIBRILLATION',  'AFIB' | 'ATRIAL FIBRILLATION HAS  REVERTED', 'ATRIAL  FIBRILLATION IS ABSENT', "IS  NO LONGER SEEN",  "REPLACES ATRIAL  FIBRILLATION", "ATRIAL  FIBRILLATION IS NO  LONGER", "REPLACED  ATRIAL FIBRILLATION",  "CHANGE FROM ATRIAL  FIBRILLATION TO", "ATRIAL RHYTHM ON THE PRIOR  EKG", "ATRIAL FIBRILLATION  WAS PRESENT ON", "[NOW  ABSENT]", "ATRIAL  FIBRILLATION ->", "ATRIAL  FIBRILLATION IS NO LONGER  PRESENT", "WAS PRESENT  ON THE PRIOR EKG" |
| QT prolongation | 'PROLONGED QT INTERVAL',  'QT INTERVAL HAS  INCREASED' | '[NOW ABSENT]' |

**Supplementary Table 2.** Reconstruction errors across different leads with different metrics for ECG12-PerceptNet and EK-GAN generator networks.

|  |  | UNet-Perceptual | |  |  | GAN | |  |
| --- | --- | --- | --- | --- | --- | --- | --- | --- |
|  | MSE  Mean(std) | MAE  Mean(std) | MMD  (10^-4) | SSIM | MSE  Mean(std) | MAE  Mean(std) | MMD  (10^-4) | SSIM |
| I | 0.33(0.35) | 0.32(0.16) | 2.82 | 0.48 | 0.47(0.39) | 0.38(0.15) | 2.82 | 0.41 |
| II | 0.34(0.36) | 0.35(0.18) | 2.82 | 0.49 | 0.46(0.39) | 0.41(0.18) | 2.82 | 0.45 |
| III | 0.12(0.12) | 0.19(0.09) | 18.07 | 0.41 | 0.15(0.13) | 0.22(0.09) | 2.85 | 0.28 |
| aVR | 0.05(0.05) | 0.14(0.06) | 2.86 | 0.49 | 0.08(0.06) | 0.17(0.07) | 2.83 | 0.44 |
| aVF | 0.04(0.04) | 0.11(0.05) | 3.03 | 0.49 | 0.07(0.05) | 0.14(0.05) | 3.41 | 0.38 |
| aVL | 0.05(0.05) | 0.14(0.06) | 2.83 | 0.51 | 0.08(0.06) | 0.17(0.06) | 2.83 | 0.40 |
| V1 | 0.35(0.42) | 0.34(0.18) | 2.83 | 0.52 | 0.52(0.60) | 0.39(0.20) | 2.83 | 0.46 |
| V2 | 0.50(0.41) | 0.40(0.15) | 2.82 | 0.67 | 0.75(0.69) | 0.47(0.19) | 2.83 | 0.58 |
| V3 | 0.50(0.40) | 0.40(0.15) | 2.82 | 0.63 | 0.73(0.70) | 0.47(0.19) | 2.82 | 0.56 |
| V4 | 0.50(0.48) | 0.41(0.18) | 2.82 | 0.58 | 0.65(0.58) | 0.47(0.19) | 2.82 | 0.54 |
| V5 | 0.47(0.47) | 0.38(0.18) | 2.82 | 0.53 | 0.61(0.53) | 0.44(0.18) | 2.82 | 0.49 |
| V6 | 0.42(0.50) | 0.36(0.20) | 2.82 | 0.45 | 0.59(0.56) | 0.43(0.20) | 2.82 | 0.41 |

Abbreviations. GAN; Generative Adversarial Network, MAE; Mean Absolute Error, MSE; Mean Squared Error, MMD; Maximum Mean Discrepancy, SSIM; Structural Similarity.

**Supplementary Table 3.** Classification performance of original and reconstructed 12lead ECG signals in test dataset (~7K ECG signals) over 5 diseases.

| Disease | Features | AUC  (95%  CI) | Accuracy (95% CI) | F1score  (95%  CI) | Precision (95% CI) | Sensitivity (95% CI) | Specificity (95% CI) | NPV (95%  CI) |
| --- | --- | --- | --- | --- | --- | --- | --- | --- |
| RBBB | Original 12-lead | 0.99  (0.99 -  0.99) | 0.96 (0.95  - 0.97) | 0.92  (0.90 -  0.93) | 0.87 (0.85  - 0.90) | 0.96 (0.95  - 0.97) | 0.96 (0.95  - 0.97) | 0.99 (0.99  - 0.99) |
|  | Reconstructed 12-lead | 0.91  (0.90 -  0.93) | 0.85 (0.83  - 0.87) | 0.71  (0.67 -  0.74) | 0.61 (0.57  - 0.64) | 0.85 (0.83  - 0.87) | 0.85 (0.83  - 0.87) | 0.95 (0.95  - 0.96) |
|  | Reconstructed (GAN) | 0.79  (0.77 -  0.81) | 0.72 (0.70  - 0.74) | 0.53  (0.50 -  0.56) | 0.41 (0.38  - 0.45) | 0.72 (0.70  - 0.74) | 0.72 (0.70  - 0.74) | 0.90 (0.89  - 0.92) |
|  | Single simulated ILR lead | 0.53  (0.51 -  0.56) | 0.50 (0.48  - 0.52) | 0.30  (0.28 -  0.33) | 0.22 (0.20  - 0.24) | 0.50 (0.48  - 0.52) | 0.50 (0.48  - 0.52) | 0.79 (0.77  - 0.81) |
|  | Single ILR lead (dedicated single-lead classifier) | 0.88  (0.87 -  0.90) | 0.81 (0.79  - 0.83) | 0.65  (0.62 -  0.68) | 0.54 (0.50  - 0.58) | 0.81 (0.79  - 0.83) | 0.81 (0.79  - 0.83) | 0.94 (0.93  - 0.95) |
| LBBB | Original 12-lead | 0.99  (0.99 -  1.00) | 0.97 (0.96  - 0.98) | 0.87  (0.84 -  0.90) | 0.80 (0.74  - 0.84) | 0.97 (0.96  - 0.98) | 0.97 (0.96  - 0.98) | 1.00 (1.00  - 1.00) |
|  | Reconstructed 12-lead | 0.94  (0.92 -  0.95) | 0.86 (0.84  - 0.88) | 0.55  (0.50 -  0.60) | 0.40 (0.35  - 0.46) | 0.86 (0.83  - 0.88) | 0.86 (0.84  - 0.88) | 0.98 (0.98  - 0.99) |
|  | Reconstructed (GAN) | 0.91  (0.90 -  0.93) | 0.83 (0.81  - 0.85) | 0.50  (0.45 -  0.54) | 0.36 (0.31  - 0.40) | 0.83 (0.81  - 0.85) | 0.83 (0.81  - 0.85) | 0.98 (0.97  - 0.98) |
|  | Single simulated ILR lead | 0.79  (0.77 -  0.82) | 0.71 (0.69  - 0.74) | 0.33  (0.30 -  0.37) | 0.22 (0.19  - 0.25) | 0.71 (0.69  - 0.74) | 0.71 (0.69  - 0.74) | 0.96 (0.95  - 0.96) |
|  | Single ILR lead (dedicated single-lead classifier) | 0.95  (0.93 -  0.96) | 0.87 (0.85  - 0.90) | 0.57  (0.51 -  0.63) | 0.43 (0.37  - 0.49) | 0.87 (0.84  - 0.89) | 0.87 (0.85  - 0.90) | 0.98 (0.98  - 0.99) |
| Prolonged QT interval | Original 12-lead | 0.88  (0.86 -  0.90) | 0.79 (0.77  - 0.82) | 0.50  (0.47 -  0.55) | 0.37 (0.33  - 0.41) | 0.79 (0.77  - 0.81) | 0.79 (0.77  - 0.82) | 0.96 (0.95  - 0.97) |
|  | Reconstructed 12-lead | 0.79  (0.77 -  0.81) | 0.72 (0.70  - 0.74) | 0.41  (0.38 -  0.44) | 0.29 (0.26  - 0.32) | 0.72 (0.70  - 0.74) | 0.72 (0.70  - 0.74) | 0.94 (0.93  - 0.95) |
|  | Reconstructed (GAN) | 0.76  (0.73 -  0.78) | 0.69 (0.66  - 0.71) | 0.37  (0.34 -  0.41) | 0.25 (0.23  - 0.28) | 0.68 (0.66  - 0.71) | 0.69 (0.66  - 0.71) | 0.93 (0.92  - 0.94) |
|  | Single simulated ILR lead | 0.67  (0.64 -  0.70) | 0.63 (0.60  - 0.66) | 0.31  (0.28 -  0.35) | 0.21 (0.18  - 0.24) | 0.63 (0.60  - 0.66) | 0.63 (0.60  - 0.66) | 0.91 (0.90  - 0.93) |
|  | Single ILR lead (dedicated single-lead classifier) | 0.67  (0.65 -  0.70) | 0.63 (0.60  - 0.66) | 0.31  (0.28 -  0.35) | 0.21 (0.19  - 0.23) | 0.63 (0.60  - 0.65) | 0.63 (0.60  - 0.66) | 0.92 (0.90  - 0.93) |
| AF | Original 12-lead | 0.99  (0.99 -  0.99) | 0.96 (0.95  - 0.96) | 0.91  (0.89 -  0.93) | 0.87 (0.84  - 0.89) | 0.96 (0.95  - 0.96) | 0.96 (0.95  - 0.97) | 0.99 (0.98  - 0.99) |
|  | Reconstructed 12-lead | 0.92  (0.91 -  0.93) | 0.86 (0.84  - 0.87) | 0.73  (0.70 -  0.75) | 0.63 (0.60  - 0.66) | 0.85 (0.84  - 0.87) | 0.86 (0.84  - 0.87) | 0.95 (0.95  - 0.96) |
|  | Reconstructed (GAN) | 0.91  (0.90 -  0.92) | 0.84 (0.82  - 0.85) | 0.70  (0.67 -  0.73) | 0.60 (0.57  - 0.63) | 0.84 (0.82  - 0.85) | 0.84 (0.82  - 0.85) | 0.95 (0.94  - 0.95) |
|  | Single simulated ILR lead | 0.89  (0.88 -  0.90) | 0.82 (0.80  - 0.83) | 0.67  (0.64 -  0.69) | 0.56 (0.53  - 0.59) | 0.82 (0.80  - 0.83) | 0.82 (0.80  - 0.83) | 0.94 (0.93  - 0.95) |
|  | Single ILR lead (dedicated single-lead classifier) | 0.85  (0.83 -  0.86) | 0.77 (0.75  - 0.78) | 0.59  (0.56 -  0.62) | 0.49 (0.45  - 0.52) | 0.77 (0.74  - 0.78) | 0.77 (0.75  - 0.78) | 0.92 (0.91  - 0.93) |
| Low EF | Original 12-lead | 0.88  (0.86 -  0.89) | 0.79 (0.77  - 0.81) | 0.47  (0.42 -  0.51) | 0.33 (0.29  - 0.37) | 0.79 (0.76  - 0.81) | 0.79 (0.77  - 0.81) | 0.97 (0.96  - 0.97) |
|  | Reconstructed 12-lead | 0.73  (0.70 -  0.75) | 0.67 (0.65  - 0.70) | 0.32  (0.29 -  0.36) | 0.21 (0.19  - 0.24) | 0.67 (0.64  - 0.70) | 0.67 (0.65  - 0.70) | 0.94 (0.93  - 0.95) |
|  | Reconstructed (GAN) | 0.67  (0.64 -  0.70) | 0.62 (0.60  - 0.65) | 0.28  (0.25 -  0.31) | 0.18 (0.16  - 0.21) | 0.62 (0.60  - 0.65) | 0.62 (0.60  - 0.65) | 0.93 (0.92  - 0.94) |
|  | Single simulated ILR lead | 0.63  (0.60 -  0.66) | 0.60 (0.57  - 0.62) | 0.26  (0.23 -  0.29) | 0.17 (0.15  - 0.19) | 0.60 (0.57  - 0.62) | 0.60 (0.57  - 0.62) | 0.92 (0.91  - 0.93) |
|  | Single ILR lead (dedicated single-lead classifier) | 0.65  (0.62 -  0.68) | 0.61 (0.58  - 0.63) | 0.27  (0.24 -  0.30) | 0.17 (0.15  - 0.19) | 0.61 (0.58  - 0.63) | 0.61 (0.58  - 0.63) | 0.92 (0.91  - 0.93) |

Abbreviations. AF; Atrial Fibrillation, AUC; Area Under Receiver Operating Curve, CI; Confidence Interval, EF; Ejection Fraction, ILR; Implantable Loop Recorder, LBBB; Left Bundle Branch Block, NPV; Negative Predictive Value, PVC; Premature Ventricular Contractions, RBBB; Right Bundle Branch Block.

**Supplementary Table 4.** Classification performance of original and reconstructed 12lead ECG signals in external test dataset (~10K ECG signals) over 5 diseases.

| Disease | Features | AUC  (95%  CI) | Accuracy (95% CI) | F1score  (95%  CI) | Precision (95% CI) | Sensitivity (95% CI) | Specificity (95% CI) | NPV (95%  CI) |
| --- | --- | --- | --- | --- | --- | --- | --- | --- |
| RBBB | Original 12-lead | 0.98  (0.98 -  0.99) | 0.94 (0.94  - 0.95) | 0.79  (0.77 -  0.81) | 0.68 (0.65  - 0.71) | 0.94 (0.94  - 0.95) | 0.94 (0.94  - 0.95) | 0.99 (0.99  - 0.99) |
|  | Reconstructed 12-lead | 0.84  (0.83 -  0.86) | 0.77 (0.75  - 0.78) | 0.43  (0.40 -  0.45) | 0.30 (0.27  - 0.32) | 0.77 (0.75  - 0.78) | 0.77 (0.75  - 0.78) | 0.96 (0.96  - 0.97) |
|  | Reconstructed (GAN) | 0.79  (0.77 -  0.80) | 0.71 (0.70  - 0.73) | 0.36  (0.34 -  0.38) | 0.24 (0.22  - 0.26) | 0.71 (0.70  - 0.73) | 0.71 (0.70  - 0.73) | 0.95 (0.95  - 0.96) |
|  | Single simulated ILR lead | 0.45  (0.43 -  0.46) | 0.45 (0.43  - 0.46) | 0.15  (0.14 -  0.16) | 0.09 (0.09  - 0.10) | 0.45 (0.43  - 0.46) | 0.45 (0.43  - 0.46) | 0.86 (0.85  - 0.87) |
| LBBB | Original 12-lead | 0.98  (0.98 -  0.99) | 0.94 (0.93  - 0.95) | 0.51  (0.46 -  0.58) | 0.35 (0.31  - 0.42) | 0.94 (0.92  - 0.95) | 0.94 (0.93  - 0.95) | 1.00 (1.00  - 1.00) |
|  | Reconstructed 12-lead | 0.93  (0.92 -  0.94) | 0.87 (0.86  - 0.89) | 0.33  (0.28 -  0.37) | 0.20 (0.17  - 0.23) | 0.87 (0.85  - 0.89) | 0.87 (0.86  - 0.89) | 0.99 (0.99  - 1.00) |
|  | Reconstructed (GAN) | 0.90  (0.88 -  0.91) | 0.82 (0.81  - 0.84) | 0.24  (0.22 -  0.27) | 0.14 (0.12  - 0.16) | 0.82 (0.80  - 0.84) | 0.82 (0.81  - 0.84) | 0.99 (0.99  - 0.99) |
|  | Single simulated ILR lead | 0.78  (0.76 -  0.80) | 0.71 (0.69  - 0.73) | 0.15  (0.13 -  0.16) | 0.08 (0.07  - 0.09) | 0.71 (0.69  - 0.73) | 0.71 (0.69  - 0.73) | 0.99 (0.98  - 0.99) |
| Prolonged QT interval | Original 12-lead | 0.79  (0.77 -  0.81) | 0.71 (0.70  - 0.73) | 0.23  (0.21 -  0.25) | 0.14 (0.12  - 0.15) | 0.71 (0.70  - 0.73) | 0.71 (0.70  - 0.73) | 0.97 (0.97  - 0.98) |
|  | Reconstructed 12-lead | 0.68  (0.66 -  0.70) | 0.62 (0.60  - 0.65) | 0.17  (0.15 -  0.18) | 0.10 (0.09  - 0.11) | 0.62 (0.60  - 0.65) | 0.62 (0.60  - 0.65) | 0.96 (0.96  - 0.97) |
|  | Reconstructed (GAN) | 0.72  (0.70 -  0.74) | 0.65 (0.64  - 0.67) | 0.19  (0.17 -  0.20) | 0.11 (0.10  - 0.12) | 0.65 (0.63  - 0.67) | 0.65 (0.64  - 0.67) | 0.97 (0.96  - 0.97) |
|  | Single simulated ILR lead | 0.64  (0.62 -  0.66) | 0.60 (0.58  - 0.61) | 0.15  (0.14 -  0.16) | 0.09 (0.08  - 0.09) | 0.60 (0.58  - 0.61) | 0.60 (0.58  - 0.61) | 0.96 (0.95  - 0.96) |
| AF | Original 12-lead | 0.97  (0.97 -  0.98) | 0.92 (0.92  - 0.93) | 0.70  (0.68 -  0.73) | 0.57 (0.54  - 0.60) | 0.92 (0.91  - 0.93) | 0.92 (0.92  - 0.93) | 0.99 (0.99  - 0.99) |
|  | Reconstructed 12-lead | 0.90  (0.89 -  0.90) | 0.82 (0.81  - 0.83) | 0.48  (0.46 -  0.50) | 0.34 (0.32  - 0.36) | 0.82 (0.81  - 0.83) | 0.82 (0.82  - 0.83) | 0.98 (0.98  - 0.98) |
|  | Reconstructed (GAN) | 0.88  (0.87 -  0.89) | 0.81 (0.79  - 0.82) | 0.45  (0.43 -  0.47) | 0.31 (0.29  - 0.33) | 0.80 (0.79  - 0.82) | 0.81 (0.79  - 0.82) | 0.97 (0.97  - 0.98) |
|  | Single simulated ILR lead | 0.83  (0.81 -  0.84) | 0.75 (0.73  - 0.77) | 0.37  (0.35 -  0.39) | 0.24 (0.23  - 0.27) | 0.75 (0.73  - 0.77) | 0.75 (0.73  - 0.77) | 0.96 (0.96  - 0.97) |
| Low EF | Original 12-lead | 0.89  (0.88 -  0.90) | 0.81 (0.80  - 0.82) | 0.41  (0.38 -  0.44) | 0.27 (0.25  - 0.30) | 0.81 (0.80  - 0.82) | 0.81 (0.80  - 0.82) | 0.98 (0.98  - 0.98) |
|  | Reconstructed 12-lead | 0.76  (0.74 -  0.78) | 0.69 (0.67  - 0.70) | 0.27  (0.25 -  0.28) | 0.16 (0.15  - 0.18) | 0.69 (0.67  - 0.70) | 0.69 (0.67  - 0.70) | 0.96 (0.96  - 0.97) |
|  | Reconstructed (GAN) | 0.72  (0.70 -  0.74) | 0.66 (0.65  - 0.68) | 0.24  (0.22 -  0.26) | 0.15 (0.14  - 0.16) | 0.66 (0.65  - 0.68) | 0.66 (0.65  - 0.68) | 0.96 (0.95  - 0.96) |
|  | Single simulated ILR lead | 0.65  (0.63 -  0.66) | 0.60 (0.59  - 0.62) | 0.20  (0.18 -  0.21) | 0.12 (0.11  - 0.13) | 0.60 (0.59  - 0.62) | 0.60 (0.59  - 0.62) | 0.94 (0.94  - 0.95) |

Abbreviations. AF; Atrial Fibrillation, AUC; Area Under Receiver Operating Curve, CI; Confidence Interval,

EF; Ejection Fraction, ILR; Implantable Loop Recorder, LBBB; Left Bundle Branch Block, NPV; Negative Predictive Value, PVC; Premature Ventricular Contractions, RBBB; Right Bundle Branch Block.
